# Tuberculosis and HIV/AIDS Co-infection in Patients Attending Directly Observed Treatment Short-Course (DOTS) Centers in Anambra State, Nigeria

**DOI:** 10.1101/2023.09.01.23294608

**Authors:** Monique Okeke, Peter M. Eze, Adaeze E. Chukwudebelu, Chidiebere J. Nwankwo, Nchekwube K. Eze, Uchenna U. Okafor, Isaiah C. Abonyi, Eric E. Okereke, Kalu O. Obasi, Okorie A. Ede, Chika P. Ejikeugwu, Cajetan I. Ilo, Jerome O. Okafor

## Abstract

**Background and Aim(s):** This study retrospectively assessed the prevalence of TB and HIV/AIDS co-infection among patients that attended the Directly Observed Treatment Short-course (DOTS) centers in Anambra State, Southeast, Nigeria, between 2013 and 2017.

**Methods:** The study adopted a descriptive and retrospective epidemiological survey design. A total of 1443 case files of patients aged 15 to 60 who were treated in DOTS centers selected from Anambra State’s 21 Local Government Areas between 2013 and 2017 were investigated. The uniform data form, a standardized instrument used in Anambra State’s health facilities for data collection, was used to collect data from case files of all those identified as co-infected with TB and HIV/AIDS.

**Results:** The mean prevalence rate of TB and HIV/AIDS co-infection in the state during the five-year period (2013–2017) was 20%. The highest annual prevalence of TB and HIV/AIDS co-infection was recorded in 2014 (23.84%). The state’s prevalence of TB and HIV/AIDS co-infection increased dramatically from 13.17% in 2013 to 23.84% in 2014, followed by a slight downward trend to 22.80% in 2015, 20.17% in 2016, and 20.03% in 2017. In terms of gender, age, marital status, and occupation, females (59.5%), those aged 15 to 25 years (30.7%), married people (43.9%), and traders/business owners (50.7%), respectively, had the highest rates of tuberculosis and HIV/AIDS co-infection during the study period.

**Conclusion:** The findings of this study show that young people, females, married people, and traders/business owners appear to be the most vulnerable groups affected by TB and HIV/AIDS co-infection, accounting for the majority of the disease burden in the state. To address the high prevalence of TB and HIV/AIDS co-infection in the Anambra State, novel intervention and control programs should be developed and implemented, and existing intervention frameworks should be strengthened.

## 1 INTRODUCTION

Tuberculosis (TB) is caused by the bacteria *Mycobacterium tuberculosis*, which typically affects the lungs, and is spread through the air, especially when people with pulmonary TB cough, sneeze, or spit (WHO, 2022a).^1^ TB primarily affects people with reduced immunity, such as young children or people living with the human immunodeficiency virus (HIV), as well as those suffering from malnutrition, diabetes, or silicosis, and those who smoke or have substance use disorders. TB also disproportionately affects people whose health is compromised due to socioeconomic factors such as poverty, poor housing, displacement, or incarceration.^2^

People living with HIV are at increased risk of dying from TB, especially if TB is not diagnosed or is diagnosed late. HIV and TB form a lethal combination, each accelerating the progress of the other. In 2021, approximately 187 000 people died of HIV-associated TB. High-quality TB screening is critical to ensuring that people living with HIV receive timely treatment for TB disease or TB infection.^1,3^

The WHO African Region has the highest burden of HIV-associated TB, and countries in Sub-Saharan Africa are the worst affected by the twin epidemic of TB and HIV. The prevalence of TB in the region, which hitherto was reported to be declining prior HIV epidemic, is now on the rise, with Nigeria among the countries with a high burden of TB.^1,4^

TB remains a serious public health challenge in Nigeria, and the country ranks among the nations with the highest disease burden. TB has a negative impact on the country’s growth and development because it causes both direct and indirect economic losses due to increased morbidity and mortality, and because a significant proportion of those affected are in productive age groups.^5^

Several TB control programmes have been initiated in developing countries, most notably Nigeria, which has a high TB burden. They include the WHO-instituted direct observed treatment short course (DOTS) program, DOTSplus, and Stop TB, all with the mandate of executing strategies aimed at reducing the global TB burden.^5-7^

Anambra State in Nigeria is a priority setting for TB control because it contributes significantly to the country’s high TB burden, accounting for the highest prevalence in the South-East region.^8^ Adebayo et al.^5^ conducted a health facility record for the year 2016, which included 22 DOTS facilities and 1281 TB treatment enrollees. They reported a prevalence rate of HIV/TB co-infection of 24.4%, compared to 7.1% in Oyo State. Their findings revealed that TB treatment success and cure rates in Anambra State fell short of the WHO’s recommended target of 85%.

Although a few other studies have reported on HIV and TB co-infection in selected Anambra State health facilities and communities,^9-11^ there appears to be a scarcity of data on the prevalence of TB and HIV/AIDS co-infection in most Anambra State communities. This has significant implications for effective planning, efficient resource allocation, and decisions about where and when to expand prevention and control activities in the state. The purpose of this study was to determine the prevalence of TB and HIV/AIDS co-infection in patients who attended DOTS centers in Anambra State, Nigeria from 2013 to 2017.

## 2 MATERIALS AND METHODS

### 2.1 Research Design

This study was a retrospective epidemiological survey of patients’ clinical records from 2013 to 2017 to determine the prevalence of TB and HIV co-infection in Anambra State, Nigeria, during that period.

### 2.2 Study Area and Study Population

This study was carried out in Anambra State, which is in Nigeria’s South-East geopolitical zone. According to the 2006 National Population Census, Anambra State has a population of over 4,055,048 people, distributed across the state’s 21 Local Government Areas.^12^ Data from hospital records of patients aged 15 to 60 who received treatments for tuberculosis and HIV/AIDS co-infection at a DOTS center selected from each of Anambra State’s 21 Local Government Areas (LGAs) between 2013 and 2017 were collected and analyzed. A total of 1443 patients’ case files/folders were sampled.

### 2.3 Data Collection

The uniform data form, a standardized data collection tool designed specifically for TB and HIV data collection in the state’s health facilities, was used for data collection. To ensure the integrity and authenticity of the data, no modifications or changes were made to the data collection instrument. Data collected included the name of the DOTS facility providing the services, the year of the patient’s treatment, the patient’s age, gender, marital status, occupation, and others.

### 2.4 Data Analysis

Data analysis was performed using Microsoft Excel (version 16.75.2). The annual prevalence of TB and HIV/AIDS co-infection in Anambra State’s 21 LGAs from 2013 to 2017 were compared using one-way analysis of variance (ANOVA) at 95% confidence level. Indicator of statistical significance is P ≤ 0.05.

## 3 RESULTS AND DISCUSSION

Table 1 shows the distribution and prevalence of TB and HIV/AIDS co-infection in DOTS centers in health facilities in each of Anambra State’s 21 LGAs from 2013 to 2017. The highest number of cases of TB and HIV/AIDS co-infection (344, 23.8%) was recorded in 2014, followed by 329 (22.8%) in 2015, 291 (20.2%) in 2016, 289 (20.0%) in 2017, and 190 (14.2%) in 2013.

**Table 1:** TB and HIV/AIDS co-infection distribution in Anambra State’s 21 LGAs from 2013 to 2017.

| S/N | Name of LGA | Name of Facility | 2013<br>(n)* | 2014<br>(n)*† | 2015<br>(n)*† | 2016<br>(n)*† | 2017<br>(n)*† | Total<br>[n (%)] |
| --- | --- | --- | --- | --- | --- | --- | --- | --- |
| 1 | Aguata | Anglican Diocesan Hospital and Maternity Igboukwu | 9 | 21 | 10 | 10 | 17 | 67 (4.46%) |
| 2 | Anambra East | Umuleri General Hospital | 7 | 17 | 15 | 13 | 13 | 65 (4.50%) |
| 3 | Anambra West | Oroma-Etiti Primary Health Centre | 8 | 31 | 20 | 20 | 27 | 106 (7.35%) |
| 4 | Awka North | Isuaniocha Primary Health Centre | 8 | 27 | 18 | 12 | 11 | 76 (5.27%) |
| 5 | Awka South | Regina Caeli Hospital, Awka | 11 | 15 | 31 | 23 | 23 | 103 (7.14%) |
| 6 | Anaocha | St. Joseph's Hospital, Adazi-Nnukwu | 16 | 21 | 22 | 17 | 18 | 94 (6.51%) |
| 7 | Ayamelum | Omasi-Agu Primary Health Centre | 12 | 36 | 25 | 17 | 12 | 102 (7.07%) |
| 8 | Dunukofia | Ukpomili Health Center, Ifitedunu | 13 | 23 | 16 | 25 | 8 | 85 (5.89%) |
| 9 | Ekwusigo | Oraifite General Hospital | 15 | 17 | 12 | 9 | 10 | 63 (4.37%) |
| 10 | Ihiala | Our Lady of Lourdes Hospital, Ihiala | 6 | 12 | 9 | 11 | 9 | 47 (3.26%) |
| 11 | Idemili North | Immaculate Heart Hospital, Nkpor | 5 | 20 | 14 | 21 | 16 | 76 (5.27%) |
| 12 | Idemili South | Our Lady of Fatima Hospital and Maternity, Awka-Etiti | 4 | 13 | 21 | 8 | 8 | 54 (3.74%) |
| 13 | Njikoka | Enugwu Ukwu General Hospital | 4 | 12 | 21 | 14 | 21 | 72 (4.99%) |
| 14 | Nnewi North | Nnamdi Azikiwe University Teaching Hospital, Nnewi | 10 | 18 | 11 | 10 | 20 | 69 (4.78%) |
| 15 | Nnewi South | Ezinifite Primary Health Centre | 6 | 10 | 7 | 20 | 16 | 59 (4.09%) |
| 16 | Ogbaru | Osomalla Primary Health Centre | 8 | 9 | 24 | 15 | 10 | 66 (4.57%) |
| 17 | Onitsha North | St. Charles Borromeo Hospital, Onitsha | 7 | 7 | 10 | 7 | 13 | 44 (3.05%) |
| 18 | Onitsha South | St. John Anglican Primary Health Care, Fegge | 13 | 14 | 12 | 13 | 17 | 69 (4.78%) |
| 19 | Orumba North | Umunebo Primary Health Centre, Ufuma | 12 | 6 | 9 | 7 | 8 | 42 (2.91%) |
| 20 | Orumba South | Immaculate Heart Hospital, Umunze | 10 | 8 | 12 | 11 | 4 | 45 (3.12%) |
| 21 | Oyi | Awkuzu Primary Health Centre | 6 | 7 | 10 | 8 | 8 | 39 (2.70%) |
| **Total [n (%)]** |  |  | 190<br>(13.17%) | 344<br>(23.84%) | 329<br>(22.80%) | 291<br>(20.17%) | 289<br>(20.03%) | 1443 (100%) |

\*P-value = 0.0015 ( $p < 0.05$ ); †P-value = 0.4506 ( $p > 0.05$ )

Figure 2 shows the prevalence of TB and HIV/AIDS co-infection in Anambra State from 2013 to 2017, based on patient demographics. In general, 585 (40.5%) of the total study population with TB and HIV/AIDS co-infection were males, while 858 (59.5%) were females. In terms of age, marital status, and occupation, cases of tuberculosis and HIV/AIDS co-infection were highest in those aged 15 to 25 years (443, 30.7%), in married patients (633, 43.9%), and in traders/business owners (731, 50.7%).

**Figure 1:**
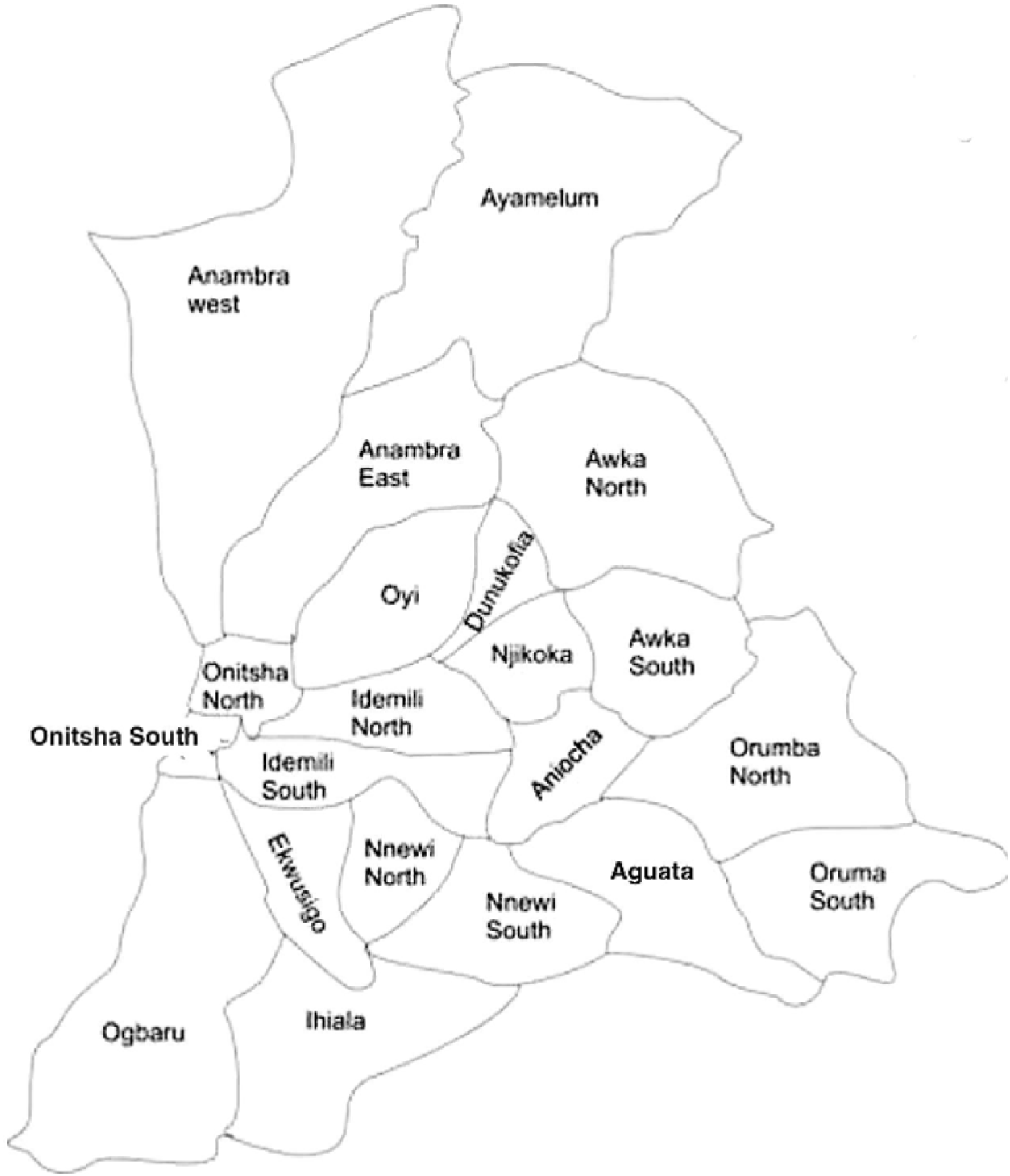
Map of Anambra State Showing the 21 Local Government Areas^13^

**Figure 2:**
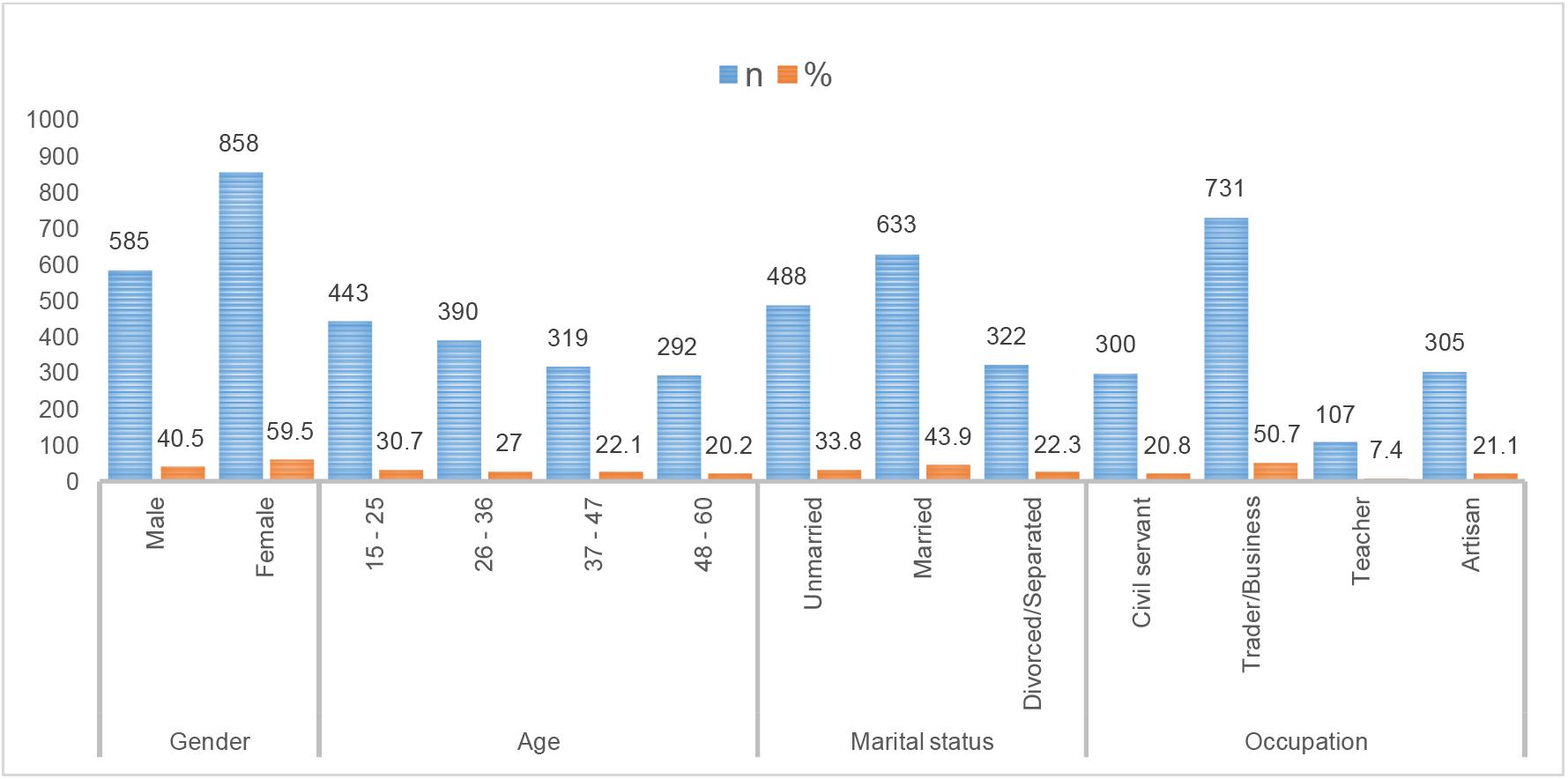
Prevalence of TB and HIV/AIDS co-infection in Anambra State from 2013 to 2017, based on patient demographics.

### 3.1 Annual Prevalence of TB and HIV/AIDS co-infection

Between 2013 and 2017, 1443 patients with TB and HIV/AIDS co-infection attended the DOTS centers sampled in Anambra State. A mean prevalence rate of TB and HIV/AIDS co-infection in the state during the five-year period (2013 – 2017) was 288.6 (20%). The highest annual prevalence of TB and HIV/AIDS co-infection was recorded in 2014 (23.84%). The state’s prevalence of TB and HIV/AIDS co-infection increased dramatically from 13.17% in 2013 to 23.84% in 2014, followed by a slight downward trend to 22.80% in 2015, 20.17% in 2016, and 20.03% in 2017.

The annual prevalence rate of TB and HIV/AIDS co-infection was significantly different (p < 0.05) over the five-year period (2013-2017), with a sharp increase from 13.17% in 2013 to 23.84% in 2014. There was no statistical difference in the annual prevalence rate of TB and HIV/AIDS co-infection from 2014 to 2017 (p > 0.05), which showed a slight downward trend of 23.84% in 2014 to 22.80% in 2015, 20.17% in 2016, and 20.03% in 2017.

Temitayo-Oboh et al.^4^ investigated the prevalence and factors influencing TB and HIV co-infection among patients attending the DOTS clinic in a tertiary health center in Ogun State, Nigeria, between 2015 and 2019. Their study showed that about one in five patients with TB had HIV infection (20% prevalence), which is consistent with our study and that another study reported in Lagos, Nigeria,^14^ and higher than the prevalence of 10.9%,^15^ 14.2%^16^ and 8.4%^6^ reported in similar studies conducted in Kano, Oyo, and Benin City, respectively.

### 3.2 Prevalence of TB and HIV/AIDS Co-infection in Relation to Demographics

From 2013 to 2017, the prevalence of TB and HIV/AIDS co-infection was higher in females (59.5%) than in males (40.5%) in Anambra State. This relative difference in the prevalence of TB and HIV/AIDS co-infection based on the gender of patients from the same environment is not surprising, as Nwobu et al.^17^ reported a similar trend of a higher prevalence rate for females over males in a comparative study of the prevalence rates of HIV among TB patients in Irrua and Benin environs of Edo State, Nigeria.

During the 5-year study period, patients aged 15 to 25 years had the highest prevalence rate of TB and HIV/AIDS co-infection (30.7%), followed by 27% for those aged 26 to 36 years, 22.1% for those aged 37–47 years, and 20.2% for those aged 48–60 years. Our study found that people aged 15 to 25 were more likely to have TB and HIV/AIDS co-infections. This age group represents a young population with the potential for poor sexual health and high-risk sexual behavior, making them particularly vulnerable to TB and HIV/AIDS co-infection.^18^ There was a general decrease in prevalence rates as patients’ ages progressed from 15 - 25 years to 48 - 60 years.

Regarding patients’ marital status, the prevalence rates for TB and HIV/AIDS co-infection cases in Anambra State during the study period were 33.8% for those who are single (unmarried), 43.9% for those who are married, and 22.3% for those who are divorced or separated. Due to the increased intensity of contact, sharing sleeping rooms, and one nursing the other, the spouse of an individual with TB and/or HIV/AIDS is highly likely to be infected.^19^ The high TB and HIV/AIDS co-infection rates among married people in this study may be due to infected married couples attending the same DOTs center, causing the records to be skewed higher for married people than for unmarried, divorced, or separated people.

In terms of occupation, traders and business owners had the highest prevalence of TB and HIV/AIDS co-infection cases (50.7%). This was followed by artisans (21.1%) and civil servants (20.8%). Teachers had the lowest percentage of cases (7.4%). Teachers’ higher literacy levels may be linked to lower TB and HIV/AIDS prevalence, as they may have a better understanding of disease prevention and control measures, and those who are already infected with TB or HIV/AIDS may follow the treatment regimen more judiciously.

### 3.3 Implications of the Findings

TB, a disease of poverty and inequality, is a leading cause of severe illness and mortality among people living with HIV. People with HIV who do not receive appropriate prevention and care are at a much higher risk of developing and dying from TB.^20^ As part of overall efforts to reduce HIV-related morbidity and mortality in high HIV prevalence settings, the WHO developed the global framework for TB/HIV with the goal of reducing TB transmission, morbidity, and mortality (while minimizing the risk of anti-TB drug resistance). This global framework largely focuses on Sub-Saharan Africa.^7^

HIV has a significant impact on TB control in countries with a high TB/HIV burden. At the same time, TB is not only the leading cause of death among people with AIDS, but it is also the most common curable infectious disease among people living with HIV/AIDS (PLWHA). As a result, it has become clear that additional interventions are urgently needed to supplement the WHO-recommended DOTS strategy for TB control.^7^

In Nigeria, there are DOTS centers in each of the 774 local government areas (districts) where TB patients can be diagnosed and treated. Some of these centers are housed within secondary and tertiary health care facilities and are staffed by community health extension workers and/or nurses who are expected to diagnose, treat, and refer patients as needed. With the implementation of the DOTS strategy in the Nigeria, treatment success rates increased from 15% in 1995 to 86% in 2017, and case detection rates increased from 4.3% to 25.8% during the same period.^5^

Data for this study were collected from hospital records of patients who received TB and HIV/AIDS co-infection treatments at a DOTS center selected from Anambra State’s 21 Local Government Areas (LGAs) between 2013 and 2017. The state’s prevalence of TB and HIV/AIDS co-infection increased dramatically from 13.17% in 2013 to 23.84% in 2014, followed by a slight downward trend to 22.80% in 2015, 20.17% in 2016, and 20.03% in 2017 (Table 1).

The sharp increase in TB and HIV/AIDS co-infection cases since 2014 is concerning, and it may reflect a failure of the state’s institutional, governmental, and community TB and AIDS control systems. This could also be due to the underperformance of the state’s existing DOTS centers, which may be overburdened due to a variety of factors, such as increased demand for their services as a result of a limited number of DOTS centers serving a large population, staff shortages, and so on.

According to Adebayo et al.,^5^ the problems of limited access to healthcare services and poor quality of DOTS centers, as well as insufficient health service capacity to deliver effective TB services that will meet the needs of patients, pose a major challenge to achieving Nigeria’s TB control targets. The negative consequences of the high prevalence of TB and HIV/AIDS co-infection cases in Anambra State are obvious. Unless and until coordinated efforts are made to control the epidemic in Anambra State, including the establishment of more DOTS centers and the expansion of the capacity of existing DOTS services, it may be difficult to achieve Nigeria’s and WHO’s goals of reducing the national and global burden of TB and HIV/AIDS.

Despite the high prevalence rate of TB and HIV/AIDS co-infection recorded in Anambra State during the study period, it is important to note that this study relied on records obtained from the state’s DOTS centers and thus did not account for cases that were not reported or treated at these DOTS centers. These unrecorded patients may be those who, for some reason, did not visit or were unaware of the DOTS centers, or who sought alternative methods of treatment elsewhere. As a result, the prevalence of TB and HIV/AIDS in the state could be higher than reported in this paper.

According to the findings of this study, younger people, particularly those aged 15 to 36 years, as well as females, married people, and traders/business owners, appear to be the most vulnerable groups affected by TB and HIV/AIDS co-infection, accounting for the vast majority of the disease burden in Anambra State. The implication is that unless special interventions and targeted control programs are directed to contain this growing epidemic among these groups of people, the state and even the country will lose a large proportion of its able-bodied men and women, who constitute a percentage of its workforce or manpower resources, to the menace of TB and HIV/AIDS.

### 3.4 Recommendations

More DOTS centers, as well as other TB and HIV/AIDS intervention programs and facilities, must therefore be established in many areas of the state to supplement the existing ones in serving the needs of the teeming population of people co-infected with TB and HIV/AIDS. The DOTS centers should be made more patient-friendly in order to encourage patients to trust and use the centers instead of seeking alternative remedies from unregulated sources. Furthermore, massive awareness campaigns and other targeted interventions in the control and prevention of the TB and HIV/AIDS epidemics in the state should be directed at the identified vulnerable groups affected by these diseases.

The findings of this study are expected to draw the attention of the state and the local governments of Anambra State, as well as health institutions and agencies in the state and Nigeria in general, and to prompt them to respond appropriately to the challenges posed by this endemic situation. Furthermore, it is hoped that results of this study will shed light on the pattern of distribution of TB and HIV/AIDS co-infection in Anambra State in terms of epidemiological variables such as age, gender, marital status, and occupation, which are associated with high rates of the diseases. This will provide the public with a clear understanding of the factors that may contribute to the spread of TB and HIV/AIDS, allowing them to make positive lifestyle choices and adopt positive behavioral changes toward the two endemic diseases.

## 4 CONCLUSION

Between 2013 and 2017, 1443 patients with TB and HIV/AIDS co-infection attended the DOTS centers sampled in Anambra State. During the five-year period, the state had a 20% mean prevalence rate of TB and HIV/AIDS co-infection, with the highest annual prevalence of 23.84% recorded in 2014. The most vulnerable groups affected by TB and HIV/AIDS co-infection appear to be young people aged 15 to 25, females, married people, and traders/business owners, accounting for the vast majority of disease burden in Anambra State. To address the state’s high prevalence of TB and HIV/AIDS co-infection rates, more intervention and control programs must be implemented, and those that are already in place must be made much more effective.

## Data Availability

All data produced in the present study are available upon reasonable request to the authors

## ACKNOWLEDGMENTS

The authors are thankful to the officials of the Anambra State Ministry of Health who provided access to the Directly Observed Treatment Course Service (DOTS) centers used in this study.

## CONFLICT OF INTEREST STATEMENT

**The authors declare no conflict of interest**.

## ETHICS STATEMENT

Ethical approval for this study was provided by the Ethical Board of Anambra State Ministry of Health, Nigeria (Approval number: MH/PRS/986/47).

## Notes

### Competing Interest Statement

The authors have declared no competing interest.

### Funding Statement

This study did not receive any funding

### Author Declarations

Ethical Board of Anambra State Ministry of Health gave ethical approval for this work. A copy of this document will be provided on request.

### Summary of Updates

Error in the abstract has been corrected. The referencing method has been changed. In addition, a missing reference has been included in the reference section.

